# Characterization of molecular heterogeneity of colorectal cancer among Bangladeshi population by targeted next generation sequencing

**DOI:** 10.1101/2022.11.15.22282336

**Authors:** Mohammad Kamruzzaman, Aura Rahman, Aparna R Karmakar, A B M Khurshid Alam, Kazi Monzur Kader, A B M Jamal, Maqsud Hossain

**Author notes:** **Corresponding author**: Muhammad Maqsud Hossain.

## Abstract

**Background:** Since the Sanger sequencing procedure was invented, scientists all over the world have been working to improve and further develop this cutting-edge technology. Next-generation sequencing (NGS) is a game-changing advancement and the ability to profile genetic changes in cancer in great detail has been made possible by recent advances in sequencing technology.

Several studies produced high-quality data in terms of mutation identification, particularly in the areas of actionable or rarely mutated genes, epigenetics, and transcriptomics. The goal of this project is to look at genetic heterogeneity in Bangladeshi colorectal cancer patients so that cost-effective precision medicine can be developed applied in Bangladesh.

**Methods:** The mutational characteristics of 24 colorectal tumor samples from Bangladeshi patients were investigated using targeted next generation sequencing to explore the association of mutations with clinical aspects. A total of 50 genes were sequenced with an average target depth of 675X.

**Results:** The left-sided tumors had higher mutation frequencies in TP53, APC, and ATM, while the right-sided tumors had higher mutation frequencies in EZH2, HNF1A, HRAS, and PIK3CA. Almost all of the samples in this study had a mutation in FGFR3. Deleterious mutations in the APC gene (94%) provided a selective advantage to the nascent intestinal tumor cell via constitutional stimulation of the Wnt signal transduction pathway and chromosomal instability. CDKN2A nonsynonymous mutations were found in almost all patient samples, with the exception of one. In a different patient with three unique mutations, 17 samples (71%) had a mutation in SMAD4 located in exon5 (p.P223N) synonymous and nonsynonymous mutations (two in NGRI04, and one in NGRI10). Several genes, including ATM, CDH1, ERBB4, and FBXW7, have been identified as being significantly affected by TNM stages.

Except for a few key colorectal cancer genes, a comparison of mutational characteristics with subjects from other countries’ independent data revealed that the identified mutational characteristics are largely Bangladesh-specific.

**Conclusion:** In Bangladesh, precision medicine development and use in colorectal cancer are poorly addressed. The pilot study will serve as a starting point for genomics research into colon cancer in poor-resource settings like Bangladesh. The relationship between metastasis and genomic markers will aid in the identification of prospective intervention markers and the development of therapeutic options at the individual level, resulting in the development of precision medicine.

## Introduction

Cancer is a leading cause of morbidity and mortality accounting for approximately 14 million new incidences and 8.2 million deaths per year [1]. Colorectal cancer (CRC) is the third most diagnosed cancer contributing to ∼1.8 million cases worldwide [1]. Despite broad screening programs, 25% of CRC patients are detected to be metastatic at initial diagnosis with one out of two diagnosed patients developing metastasis [2]. Prevalence studies of CRC among Bangladeshi population over the last five years have reported a total of 10,000 cases with ∼5,000 new cases every year and mortality as high as ∼4300 (∼4.4%) [1].

CRC exhibits tremendous heterogeneity with spatially and temporally observable differences in genetic mutation, epigenetic regulation and tumor microenvironment [3]. Understanding the genetic variations within a given tumor and between different metastatic sites of the tumor can yield better understanding of tumorigenesis, tumor development and progression. Insights from such studies will have immense clinical implications leading to more informed treatment decisions being implemented.

Next Generation Sequencing (NGS) has proven to be a powerful tool in detecting millions of somatic mutations and genetic alterations in cancer cells [4]. The falling costs and greater accessibility to NGS facilities in recent years have led to a shift in molecular technologies used in research, with NGS gaining rapid popularity. With advancements in NGS technologies over the years, a large section of cancer research has focused on uncovering the genetic basis of cancer development and progression using targeted NGS techniques. A key challenge in studying cancer genomes is determining which genetic mutations drive cancer progression. Frequency-based and function-based approaches have thus been developed to identify candidate markers [5].

Recent advancements in targeted therapy and personalized medicine have led to increased enthusiasm in incorporating genetic profiling of tumors into regular practices of cancer screening and treatment [6]. Almost daily, new information surfaces regarding the role of different molecular biomarkers in cancer prognosis and their theranostic significance. This raises the need for validated, sensitive and widely available molecular screening tests for implementation and improvement of multi-modal treatment strategies and clinical trials.

In the present study, we sought to determine genetic heterogeneity of CRC patients among Bangladeshi population using targeted genome sequencing by the latest NGS technology. Findings from this study will aid in the development of precision therapy for CRC patients. This pilot study will also lay the groundwork for the discovery of novel biomarkers with clear diagnostic, therapeutic, or prognostic value for individual patient monitoring at a low cost.

## Methods of the Study

### Sample collection

A total of 24 samples with colorectal cancer were obtained from the National Institute of Cancer Research & Hospitals (NICRH) from July 2022 to October 2022. Samples were obtained from formalin-fixed, paraffin-embedded archived surgical specimens and/or blood specimens.

### Factors in Study (variables)

Age, sex, family history, stage of tumor, histological grading, etiological factors, smoking, alcohol intake, DNA quality and characteristics, genome variation (compared to variables available from online databases such as TCGA, dbSNP, ClinVar etc. This will be an observational study with applied Genomics and Bioinformatics.

### Study population

All patients had to personally sign a written consent before embarking in the study and the study has to be approved by the ethical committee of Bangladesh Medical Research Council (BMRC).

#### Inclusion criteria

a. Good performance status.
b. Age 20-70 years.
c. Histologically proven colorectal cancer patients with good performance status.
d. Tumor stage I–IV.

#### Exclusion criteria

a. Moribund patients.
b. Life expectancy < 1 year.
c. Inability or refusal to perform informed consent.
d. Inability to comply with the control or intense follow-up program.

### DNA isolation

The Formalin-fixed, paraffin-embedded (FFPE) specimens will be subjected to histological review, only those containing sufficient tumor cells (at least 70% tumor cells), as determined by hematoxylin and eosin staining, will be selected for DNA isolation. Genomic DNA will be isolated by using the QIAamp1 DNA Mini Kit (Qiagen, Hilden, Germany) according to the manufacturer’s procedure with slight modifications. Genomic DNA samples will be quantified by using the NanoDrop 2000 spectrophotometer (Agilent, Santa Clara, CA, USA). The isolated DNA was stored at −80°C until analysis.

### Target enrichment and sequencing

Sample sequence enrichment and library preparation was conducted by using the SureSelect Target Enrichment Kit (Agilent, Santa Clara, CA, USA) and AmpliSeq Cancer hotspot panel (illiumina) according to the manufacturer’s instructions. The resulting purified libraries were pooled and sequenced by using the Illumina1 MiSeq™ NGS instrument for 2 × 150 paired-end sequencing reads (Illumina, San Diego, CA, USA) according to the manufacturer’s protocols. The genes in the cancer panel are listed in S1 Table. Bioinformatics analysis Sequencing data were accessed through the MiSeq Reporter. Data quality was checked by using FastQC (http://www.bioinformatics.bbsrc.ac.uk/projects/fastqc/) and then aligned to the human reference genome (hg19) by using the Burrows–Wheeler alignment tool to generate a bam file [11]. Variant calling was conducted by using the Genome Analysis Toolkit, by default [12]. Raw variant calls were filtered out by using allele frequencies > 5% and altered reads>7X. Then, the resulting variants were annotated by SNPeff and ANNOVAR [13]. Known single-nucleotide polymorphisms were excluded by using variants in the dbSNP 137 (hg19) [14] and SNPs presented in the 1000 Genomes data. All the mutational analysis are provided in supplementary File 1.

### Data Interpretation and Statistical Analysis

Clinical characteristics including age, sex, and ancestry were used in regression models to control for differences between cases and controls. For tests of genetic heterogeneity, we used the Breslow–Day test. All analyses were done using the program PLINK and SPSS. Clinical characteristics were compared between different variants (data from dbSNP and The Cancer Genome Atlas).

### Ethical Declaration

All relevant ethical guidelines have been followed, and the study was approved by North South University IRB / ethics committee approvals have been obtained.

## Results

### Clinical characteristics of patients

A total of 24 samples with colorectal cancer was obtained from National Institute of Cancer Research & Hospitals (NICRH) from July 2022 to October 2022. For two cases, we received poor sequencing data and were excluded from further mutational analysis. Patients’ characteristics are described in Table 1. Among the patients are xx male (%) and female (%), and age distribution shows that most of the patients (n=14, 51.9%) were over 50 years of age while 8 (29.6%) were between 30-50 and 2 (7.4%) were below 30 years old. Eleven (44.83%) patients had the smoking habit among the 24 patients. According to anatomical classification, 15 (55.6%) were classified as left hemicolon carcinoma and other 9 (33.3%) patients were right hemicolon carcinoma. The staging of cancers describes the spread of cancer in the body. According to the American Joint Committee on Cancer (AJCC) system, the earliest colorectal cancers are called state 0 (a very early cancer), then from stagesI (1) through IV (4). The lower the number, the less the cancer has spread. Among the 30 samples the distribution based on TNM staging was stageI (n=3, 12.5%), stageII (n=7, 29.17%), stageIII (n=9, 37.5%), and stageIV (n=3, 12.5%), respectively. Apart from TNM staging system, histological classification is another important factor influencing the clinical features and outcome of CRC patients, and we found most of the patients were classified as histological grade II (n=16, 66.67%), while 4 samples (16.67%) were found in each groups, histological gradeI and gradeIII. Patients were categorized three groups, low (<2.5 ng/ml, n=11, 45.83%), high (high >=2.5, n=4, ng/ml), and elevated (>=5 ng/ml, n=9, 37.5%) according to preoperative serum CEA level.

**Table 1:** Clinical characteristics of patients with colorectal cancer

| Characteristics |  | Frequency | Proportion (%) |
| --- | --- | --- | --- |
| **Age** | <30 years | 2 | 2 |
|  | 30-50 years | 8 | 8 |
|  | >50 years | 14 | 14 |
| **Smoking habit** | yes | 11 | 40.7 |
|  | no | 13 | 48.1 |
| **Sex** | Male | 17 | 71 |
|  | Female | 7 | 29 |
| **Presentation with acute abdomen** |  |  |  |
|  | yes | 5 | 20.83 |
|  | no | 19 | 79.17 |
| **Smoking** |  |  |  |
|  | yes | 11 | 45.83 |
|  | no | 13 | 54.17 |
| **Red Meat (overcooked)** |  |  |  |
|  | yes | 11 | 45.83 |
|  | no | 13 | 54.17 |
| **Types of treating Hospital** |  |  |  |
|  | Capital city private | 17 | 70.83 |
|  | Urban Private | 3 | 12.50 |
|  | Capital City Public | 4 | 16.67 |

| SITE OF COLORECTAL<br>CANCER |  |  |  |
| --- | --- | --- | --- |
|  | Right Colon | 9 | 37.50 |
|  | Left Colon (includes<br>Rectum) | 15 | 62.50 |
| TNM staging |  |  |  |
|  | TNM StageI | 3 | 12.50 |
|  | TNM StageII | 7 | 29.17 |
|  | TNM StageIII | 9 | 37.50 |
|  | TNM Stage IV | 3 | 12.50 |
| Histologic grade |  |  |  |
|  | Histological Grade 1 | 4 | 16.67 |
|  | Histological Grade 2 | 16 | 66.67 |
|  | Histological Grade 3 | 4 | 16.67 |
| Preoperative CEA |  |  |  |
|  | Low (<2.5ng/ml) | 11 | 45.83 |
|  | High (>=2.5 ng/ml) | 4 | 16.67 |
|  | Elevated (>=5.0 ng/ml) | 9 | 37.50 |
| Family history |  |  |  |
|  | yes | 3 | 12.50 |
|  | no | 21 | 87.50 |

### Mutational landscape of exonic regions in Bangladeshi Colorectal Cancer Patients

We sequenced 50 cancer-related genes in 24 colorectal tumors, however, due to poor sequence quality 20 samples were considered for further analysis. The mean read depth of sequencing was 650.5x per gene. Different types of mutations in coding regions including synonymous, missense, frameshift, and non-coding regions such as 5’- and 3’-UTR were found (Figure 1). Mutational frequencies were higher in non-coding regions compared to coding regions as anticipated and we found fewer non-synonymous mutations in exons than expected from their sequence content and demonstrating that such mutations are not due to purifying selection. We further investigated mutational burden only in exonic regions.

**Figure 1:**
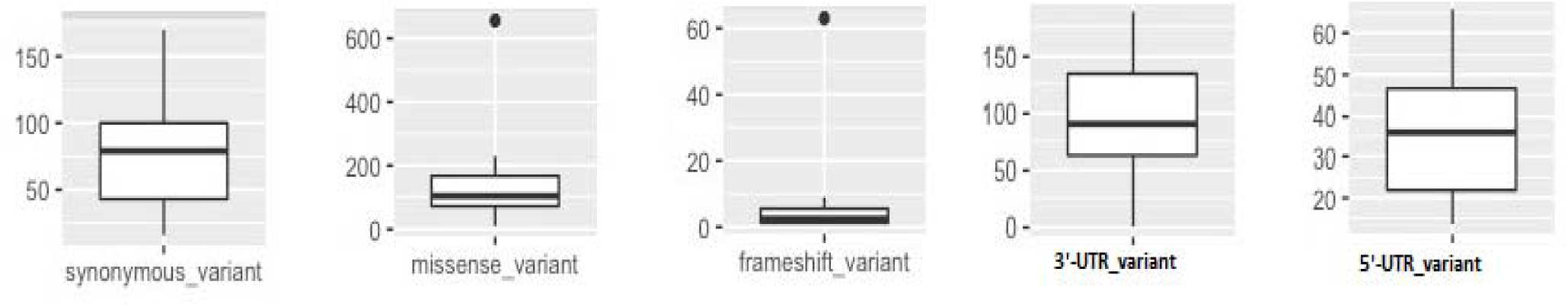
Different types of mutational frequencies in 20 samples.

To examine the clinical utility to apply targeted sequencing in the clinical settings the illumine cancer hotspot panel was used to identify mutations in 50 oncogenes and tumor suppressor genes in 22 colorectal cancer samples (two samples excluded due to poor sequencing quality) from Bangladeshi patients.

Gene-wise mutational frequency is provided in Supplementary File 1. Analysis revealed that all samples had a synonymous mutation in gene FGFR3 and 21 of 22 samples (95.5%) contained frequent mutations in genes APC, CDK2NA, MPL, RET including PDGFRA (20, 91%), ERBB4 (21, 87%), KDR (21, 87%), RB1, STK11, FLT3, and KIT (18, 82%) and among the other known CRC causing genes TP53(n=16, 72%), PIK3CA (n=2, 8.3%) and less frequent mutations in (less than 10% more than 2%) KIT, ERBB2, KIT, MPL, FBXW7, MET, and PTEN. There was no significant mutations were found between female and male for any of the genes except TP53 (65% vs 34.65%) and PIK3CA (84.6% vs 15.4%).

All the samples had a synonymous mutation in gene FGFR3 (pos. chr4:1806167; exon11:c.G1617A:P.T539T)) - fibroblast growth factor receptor 3 and 23 out of 24 patients had nonsynonymous mutations in position chr4:1801892:exon3:c.T257G:p.V86G. Four of our 22 samples (18%) had one or more mutations in the Wnt signalling pathway.

APC mutations were found in several positions and the synonymous mutations were found in 21 samples (95.5%) another frameshift mutation was found in two samples while unique mutations were found in while 7 other unique mutations were found and among these mutations 6 were harboring nonsynonymous/frameshift mutations, implicating the role of somatic mutation in APC genes in colorectal cancer.

CDKN2A (chr9:2191138, exon1:c.A68C: p.D23A) nonsynonymous mutation was found in 21 samples, followed by the nonsynonymous mutation (exon1:c.A122C:p.D41A) in 17 patients (77%), and another nonsynonymous mutation (pos. chr9:21971234). One synonymous mutation (exon1: cA268T) and another synonymous mutation (exon2:c231T) were found unique to patients NGRI04 and NGRI15, respectively.

Nonsynonymous mutations in gene MPL proto-oncogene, thrombopoietin receptor (chr1: 43349400: c.T606G:p. W536G) was found in in 21 patients, conferring as one of the germline marker for Bangladeshi colorectal patients. Two patients had one nonsynonymous mutation and two other patients had two synonymous mutations.

In RET, one synonymous mutation was found in all patients while 1 synonymous mutation (chr10:43120185) was found in 13 patients (59%). We found four additional nonsynonymous mutations in RET gene in 3 and 1 patients, respectively.

We found only one synonymous mutation in PDGFRA in position chr4: 54274888 (c.A1701G:p.567p) in 21 patients (96%). Patients with gastrointestinal stromal tumors or myeloid malignancies who have hypereosinophilia have been found to have activating mutations in the platelet-derived growth factor (PDGF) receptor alpha (PDGFRA) [27].

Single mutations in genes STK11, FLT3 KIT, GNAS, ATM were found in 20 samples (>80% cases). However, nonsynonymous mutations a number of genes CD2KNA (exon1:c.A122C:p.D41A), FBXW7(exon8: c.A1018G: p.S340G), ERBB4(exon5: c.G479C: p.G160A), SMAD4(exon5:c.C668A:P. T223N), TP53 (exon3: c. C215G: p. P72R), PTEN (stopgain, exon2: c.A94T: p.72R) and KDR (exon7: c. T900G: p. S300R) with mutations known to play key roles in different cancer types in different studies. Mutations in these genes were found in more than 70% cases.

Sixteen of the 22 samples (73%) had a mutation in FBXW7 found at exon4 (p.S340G), exon8 (p.R465C). Seven of nine mutations found in combination with KRAS. Seventeen of the 22 (68%) had 1 or more mutations in the RAS signalling pathway which include the oncogenes BRAF, KRAS, and NRAS. The majority of RAS mutations were found in KRAS, where 10 samples (45%) harbored mutations in exon2.

Twenty of 22 samples (91%) had a mutation in PIK3CA or ERBB2 in the phosphatidylinositol 3-kinase (PIK3CA) signaling pathways 12 samples (55%) contained PIK3CA mutations found in exon 5 (p.R309T), exon 21 (n=8, 37%)) or exon 5 and eight unique mutations were found in exons, 2, 10, 21, 7, 5, and in 8).

17 of 22 (78%) samples harbored mutations in TP53, all in exon5 (p C215G), and eight unique mutations were found in exon 3, 4 and 6). Nineteen of the 20 (86%) TP53 mutations occurred in combination with other genes and nearly all (90.5%) combined with mutation in RAS pathway.

### Genomic characteristics of Bangladeshi colorectal cancers: correlation with different clinical and diagnostic outcome

We compared the mutation frequencies between 14 patients with left-sided tumors and 8 patients with right-sided tumors. When comparing the mutation frequencies of illumina cancer hotspot genes [15], TP53 (p = 0.005) and APC (p = 0.013), ATM (p=0.05), showed significant difference between left-sided and right-sided tumors with higher mutation frequencies on the left-sided tumors. In a previous study [15], TP53 also showed significantly higher mutation frequency on the left-sided tumors than the right-sided tumors, which is consistent with our result. Their results also showed the same left/right-side preference of mutation frequencies for APC, but their result did not accompany statistical significance. This slight difference between two studies may be due to the heterogeneity in genomic characteristics between subjects from Bangladeshi populations of our study and the subjects from the previous study. A number of other genes apart from genes mentioned above (known to play role in colon cancer) that showed large differences in mutation frequencies between the left-sided and right-sided tumors (Fig. 1B). BRAF (p = 0.05), which is known to play a role in cell growth by sending signals inside the cell promoting, among other functions, cell division showed a significantly higher mutation frequency from the left-sided tumors than the right-sided tumors. EZH2, HNF1A, HRAS, PIK3CA showed higher mutation frequencies from the right-sided tumors than the left-sided tumors.

Mutations in genes ABL1, ALK, ATM, BRAF, CDH1, CDKN2A, EGFR, ERBB2, ERBB4, FBXW7, FGFR1, FGFR2, FLT3, HNF1A HRAS, IDH1, MET, NRAS, PIK3CA, RB1, RET, STK11 and TP53 were found significantly different (p <0.05) between smoking and non-smoking patients. Interestingly, mutations in most of the mutations in these genes was higher in smoking groups. However, the finding needs to be tested in larger sample size, and moreover, the smoking habit is less prevalent in the female population.

We also looked for the gene, mutations in which are significantly different between patients with family history. APC, ATM, BRAF, EZH2, HNF1A, HRAS, PIK3CA and TP53 (p<0.05). Based on histological classification, a significant difference was found in genes ABL1, ATM, CDH1, ERFF2 (p < 0.05).

Mutations in genes ABL1 found in samples with histological grade 1 (n=3), histological grade 2 (n=1), ATM mutations were found only in samples with histological grade 3, CDH1 mutation was found in Grade 1, and mutation in FLT3 was found in samples with Histological Grade3. Comparison of mutation among different TNM stages identified several genes including ATM CDH1 ERBB4, and FBXW7 which are significantly different (p < 0.05) among different groups. Interestingly, all mutations were found in patients at stage IV. No association was found in regards to preoperative CEA.

## Discussion

In this study, almost all samples harbored mutation in FGFR3. Previous research has shown that FGFR3-IIIc has an oncogenic role in colorectal cancer via mediating FGF18 actions, and it could be a promising novel therapeutic target. Another findings show that a FGFR3 p.R248H mutation is involved in the carcinogenesis of a subset of Lung Cancers and with our finding it is evident that mutation in this gene may aid in the future explanation of FGFR3 mutation-positive in different cancers such as CRC [28].

Deleterious mutations were found in APC gene (94% cases) in this study. Adenomatous polyposis coli (APC) gene mutations not only cause familial adenomatous polyposis (FAP), but they also have a role in the majority of sporadic colorectal malignancies. The mutation of APC has been shown to provide a selective advantage to the nascent intestinal tumor cell through constitutional stimulation of the Wnt signal transduction pathway and chromosomal instability.

As we can see different mutations including nonsynonymous mutations were found in CDKN2A and found in almost all patient samples excluding one patient. CDKN2A (the HUGO-approved official gene symbol for p16/ink4a) is a tumor suppressor that inhibits cyclin-dependent kinase 4 (CDK4) and CDK6 [29]. In a wide range of human tissue, CDKN2A (p16) expression is increased in cellular senescence and increases dramatically with aging. 1 CDKN2A is a cancer-prevention protein that can also induce aging by causing cell growth arrest and senescence [29]. Deletion, point mutation, and/or promoter methylation of CDKN2A may result in uncontrolled cell proliferation and neoplastic transformation.

*RET* encodes a transmembrane tyrosine kinase receptor that has three isoforms, long (RET51), intermediate (RET43), and short (RET9) and the mutation in RET may lead to the activation of atypical signaling pathways, which could play an important role in cancer formation [30] [31]. MPL known a proto-oncogene was also found to be frequently mutated. According to STRING and KEGG, MPL interacts with proteins involved in cell formation, apoptosis, signal transduction, and various cancer pathways including colorectal cancer, lung cancer, pancreatic, and skin cancer. According to a previous study, Y252H has been associated to the development of essential thrombocythemia [32].

As mentioned, of the 17 samples (71%) had a mutation in SMAD4 located in exon5 (p.P223N) and exon8 (p.A392V) and occur as a relatively late event with increasing incidence found with progressive disease stage. We also found several other synonymous and nonsynonymous mutations in a different patient with 3 unique mutations (two in NGRI04, and one in NGRI10). Inactivating mutations in SMAD4 are found in 5.0–24.2 percent of colorectal tumors (CRCs), making it one of the most frequently altered genes in CRC.

Our study shows that using deep targeted sequencing for the molecular characterization of colorectal cancer can reveal various mutational patterns that can be correlated with clinical features. Although numerous genomic studies have been reported for colorectal cancer, to the best of our knowledge this is the first study which few studies explored the link between mutational characteristics and various clinical features from Bangladeshi populations using next next-generation sequencing. It has been suggested that left- and right-sided colorectal cancers differ in their associated genetic alterations in neoplastic transformation based on studies in Western countries, especially Australia and Europe [33] [34]. From our study, TP53, APC, and ATM TP53 and APC showed significantly higher mutation frequencies on the left-sided tumors than the right-sided tumors. These mutations might confer a selective advantage to carcinoma cells in the left-sided colon. A previous report [15] also showed significantly higher mutation frequency of TP53 from the left-sided tumors, but they did not report the preferential mutation of APC on the left-sided tumors with statistical significance. This discrepancy from our study in comparison with the previous report can be due to the heterogeneity in genomic characteristics from Bangladeshi population. It has been suggested that left- and right-sided colorectal cancers differ in their associated genetic alterations in neoplastic transformation based on studies in Western countries, especially Australia and Europe [33] [34].

On the other hand, EZH2, HNF1A, HRAS, and PIK3CAPRKDC and ATRX genes showed significantly higher mutation frequencies from the right-sided tumors. EZH2 expression has been found to be correlated with the adenoma-carcinoma sequence of colorectal cancer. Both of these genes can be associated with more frequent observation of highly mutated MSI-high cases with right-sided tumors, as PRKDC plays a role in DNA double strand break repair [[35].

HNF1A expression was associated with resistance to anticancer drug treatment, and its suppression improved anticancer drug sensitivity. The clinicopathologic features and frequency of *KRAS* mutations in colorectal cancer (CRC) patients are well addressed; however, the impact of *HRAS* mutations on the survival of CRC patients have seldom been addressed. In many cancers, including colorectal cancer, mutations in PIK3CA, the catalytic subunit of PI3K is well documented. As in this study, PIK3CA mutations have been detected in 10–20% of CRC cases, with about 80% of mutations in two hot regions in exon 9 and exon 20. PIK3CA mutations have been linked to a worse clinical outcome and a negative prediction of a response to anti-EGFR monoclonal antibody therapy in RAS wild-type CRC. However, not all studies have supported these findings, and more extensive study has found that these effects may be limited to Exon 20 mutations. Thus, PIK3CA mutations appear to be a risk factor.gene for the management of Bangladeshi patients with CRC [36]. The substantial molecular difference between right and left-sided colorectal cancers could be attributed to the left and right colon’s embryonic origins. Our discovery highlights the significance of addressing mutations in these genes, as well as other heavily altered genes.

Frequent mutations of genes ABL1, ERBB2, FLT3, and HRAS apart from ATM were found among different age groups. Early onset colorectal cancer patients from our study showed more frequently mutated NOTCH1 tumor suppressor and the DNA strand break repair gene PALB2, and mutations of these genes can be related to the early tumor initiation and generally worse prognosis from young colorectal cancer patients.

TNM is a system for describing the amount and spread of cancer in the body of a patient. T stands for tumor growth and any cancer spread into neighboring tissue; N stands for cancer spread to nearby lymph nodes; and M stands for metastasis (spread of cancer to other parts of the body). The American Joint Committee on Cancer (AJCC) and the International Union against Cancer (IUAC) developed and maintained this system (UICC). Identification of most of the mutations in Stage IV implies the significance of the role of genes ATM, CDH1 ERBB4, and FBXW7 in cancer progression. Mutations in the CDH1 gene are thought to cause a nonfunctional E-cadherin protein. In these cells, the absence of functional E-cadherin impairs tumor suppression and cell adhesion, resulting in fast cell proliferation and metastasis. However, in other studies advanced colorectal cancer had more frequent KRAS and PTEN mutations which is different from our studies [37]. This justifies future mutational investigation among Bangladeshi groups in order to detect mutations that are important to the indigenous population.

The clinical association mutational characteristics from the TCGA study’s Caucasian subject samples revealed low overlap, with only a few essential key colorectal cancer genes like APC and JAK1 being prevalent [37]. This shows that our findings point to colorectal cancer mutational characteristics unique to the Bangladeshi population.

There were certain limitations of the study. To begin, we did not characterize structural variations such as CNVs because targeted sequencing has only had little success in finding structural variations. Second, we did not include matched normal DNA in our analysis, which limits our capacity to discover more precise somatic mutations in cancers. We used strict criteria to eliminate germline mutations that could have been mistaken for false-positive mutations.

However, without normal DNA, there is no way to correctly identify somatic mutations, and our results may contain some level of variance. Despite these limitations, we believe that our research will contribute to a better understanding of the clinical and molecular aspects of colorectal cancer by providing information on mutational characteristics from Bangladeshi patients.

## Data Availability

Datasets will be available upon request

